# Fact Check: Assessing the Response of ChatGPT to Alzheimer’s Disease Statements with Varying Degrees of Misinformation

**DOI:** 10.1101/2023.09.04.23294917

**Authors:** Sean S. Huang, Qingyuan Song, Kimberly J. Beiting, Maria C. Duggan, Kristin Hines, Harvey Murff, Vania Leung, James Powers, T.S. Harvey, Bradley Malin, Zhijun Yin

## Abstract

**Background:** There are many myths regarding Alzheimer’s disease (AD) that have been circulated on the Internet, each exhibiting varying degrees of accuracy, inaccuracy, and misinformation. Large language models such as ChatGPT, may be a useful tool to help assess these myths for veracity and inaccuracy. However, they can induce misinformation as well. The objective of this study is to assess ChatGPT’s ability to identify and address AD myths with reliable information.

**Methods:** We conducted a cross-sectional study of clinicians’ evaluation of ChatGPT (GPT 4.0)’s responses to 20 selected AD myths. We prompted ChatGPT to express its opinion on each myth and then requested it to rephrase its explanation using a simplified language that could be more readily understood by individuals with a middle school education. We implemented a survey using Redcap to determine the degree to which clinicians agreed with the accuracy of each ChatGPT’s explanation and the degree to which the simplified rewriting was readable and retained the message of the original. We also collected their explanation on any disagreement with ChatGPT’s responses. We used five Likert-type scale with a score ranging from -2 to 2 to quantify clinicians’ agreement in each aspect of the evaluation.

**Results:** The clinicians (n=11) were generally satisfied with ChatGPT’s explanations, with a mean (SD) score of 1.0(±0.3) across the 20 myths. While ChatGPT correctly identified that all the 20 myths were inaccurate, some clinicians disagreed with its explanations on 7 of the myths.

Overall, 9 of the 11 professionals either agreed or strongly agreed that ChatGPT has the potential to provide meaningful explanations of certain myths.

**Conclusions:** The majority of surveyed healthcare professionals acknowledged the potential value of ChatGPT in mitigating AD misinformation. However, the need for more refined and detailed explanations of the disease’s mechanisms and treatments was highlighted.

**Impact Statement:** There are many statements regarding Alzheimer’s disease (AD) diagnosis, management, and treatment circulating on the Internet, each exhibiting varying degrees of accuracy, inaccuracy, and misinformation. Large language models are a popular topic currently, and many patients and caregivers may turn to LLMs such as ChatGPT to learn more about the disease. This study aims to assess ChatGPT’s ability to identify and address AD myths with reliable information. We certify that this work is novel.

**Key Points:**

- Geriatricians acknowledged the potential value of ChatGPT in mitigating misinformation in Alzheimer’s Disease
- There remain nuanced cases where ChatGPT explanations are not as refined or appropriate.
- Why does this matter? Large language models such as ChatGPT are very popular nowadays and patients and caregivers often may use them to learn about their disease. The paper seeks to determine whether ChatGPT does an appropriate job in moderating understanding of Alzheimer’s Disease myths.

## Introduction

Over 80% of American adults now use the Internet to obtain health information.^1^ This democratization has led to an increase in access to information as well as an escalation in the spread of health-related misinformation, posing severe challenges to public health and patient decision-making.^2^ Broad or lay public awareness of a health issue or a disease, including Alzheimer’s disease (AD), often coexists with certain misconceptions.^3^ In this respect, there are widely accessed AD-related statements that have been posted online, each demonstrating varying degrees of accuracy, inaccuracy, and misinformation. While aware of the disease, many patients and caregivers, whether through experience or training, lack certain knowledge regarding its diagnosis and management.^4^ Through public access to misinformation circulated and gathered from non-clinical sources, not only is AD often unfairly stigmatized but medical professionals are under consulted. This trend has led to some clinicians who treat AD to call for action to help distinguish fact from fiction.^5^

The emergence of large language models (LLMs) has created a new opportunity for people to search for relevant medical knowledge. LLMs comprise a complex neural network with billions of parameters estimated on large quantities of data. This platform and training process enables LLMs to capture and generate complex linguistic patterns and dependencies, thus supporting an ability to convey and express human-like text based on input prompts.^6^ The commonly known LLMs, such as ChatGPT (or GPT-4^7^) developed by OpenAI have shown remarkable capabilities in language understanding and contextually relevant text generation.^8-12^ However, LLMs are not infallible and do not exist outside of human culture, as such they can mirror the assumptions and prejudices inherent in their training data, leading to biases and misinformation in their responses, raising ethical and safety concerns.^13^ Thus, to leverage LLMs to combat AD misinformation, it is necessary to examine their performance through verification with domain or subject matter experts. Notably, ChatGPT has been shown to effectively recognize myths in cancer,^14^ but no study has shown its benefits in AD. Therefore, the objective of this study is to assess ChatGPT’s ability to accurately address AD myths and provide reliable information.

## Methods

We conducted a cross-sectional study of clinicians’ evaluation of ChatGPT’s opinion of 20 AD myths (see Table 1). In this investigation, “myths” are understood as sets of ideas that become accepted and/or otherwise understood by communities according to dominant ideals that may themselves be based on varying degrees accuracies, inaccuracies, and misinformation. We explain how AD myths were selected and validated in the Supplemental Information (SI). The study took place between April 2023 and May 2023 and was deemed to not be human subjects research by the Vanderbilt University Medical Center Internal Review Board.

**Table 1.** The 20 Alzheimer’s disease myths used in this study.

| Index | Myth |
| --- | --- |
| 1 | Alzheimer's disease symptoms are a normal part of aging |
| 2 | Symptoms of memory loss must be Alzheimer's disease |
| 3 | All types of dementia are Alzheimer's disease |
| 4 | All Alzheimer's disease patients are disoriented, look confused, and act outside social norms |
| 5 | Alzheimer's disease is a shameful diagnosis |
| 6 | An at-home genetic test can tell me if I have (or will have) Alzheimer's disease |
| 7 | Alzheimer's disease is not fatal |
| 8 | Alzheimer's disease patients are close to death |
| 9 | Alzheimer's disease patients cannot get depressed |
| 10 | I will develop Alzheimer's disease if my parent has it |
| 11 | Only older people get Alzheimer's disease |
| 12 | Supplements found online can prevent or cure Alzheimer's disease |
| 13 | Specific diets can reduce the risk of Alzheimer's disease |
| 14 | Prescription medications can reverse the progression to Alzheimer's disease |
| 15 | Non-steroidal anti-inflammatory drugs (NSAIDs) can prevent Alzheimer's disease |
| 16 | Ginkgo biloba can prevent Alzheimer's disease |
| 17 | Hormone therapy prevents and treats Alzheimer's disease |
| 18 | Vaccines increase the risk of Alzheimer's disease |
| 19 | Aspartame increases the risk of Alzheimer's disease |
| 20 | Statins increase the risk of Alzheimer's disease |

We prompted ChatGPT to generate an opinion for each myth and then rewrite the technical explanation using non-specialist terms such that recipients with a middle school education-level could understand it. We then developed and administered a survey to inquire about the degree to which clinicians agreed with the accuracy of each original explanation and the extent to which the rewritten (simplified) explanation was readable, and retained the information in the original.

Figure 1 provides a sample of the survey questions for Myth #3. The survey begins with a “To ChatGPT:” component to inquire about ChatGPT’s opinion on the myth. Next, the survey asks the clinician if they are satisfied with ChatGPT’s opinion. The clinician could express their disagreements with the ChatGPT’s opinion through a binary ‘yes/no’ question, complemented by an optional text field to discuss the disagreements. The second “To ChatGPT:” component contains ChatGPT’s rewritten explanation and two questions for the clinicians about their assessment of its information retention and readability. In the survey, these questions use a five-point Likert-type scale that ranged from either “Very Unsatisfied” or “Strongly Disagree” to “Very Satisfied” or “Strongly Agree”. Each option was associated with an according integer score ranging from -2 to 2. As such, a positive score indicates that clinicians favor ChatGPT’s explanation or rewriting.

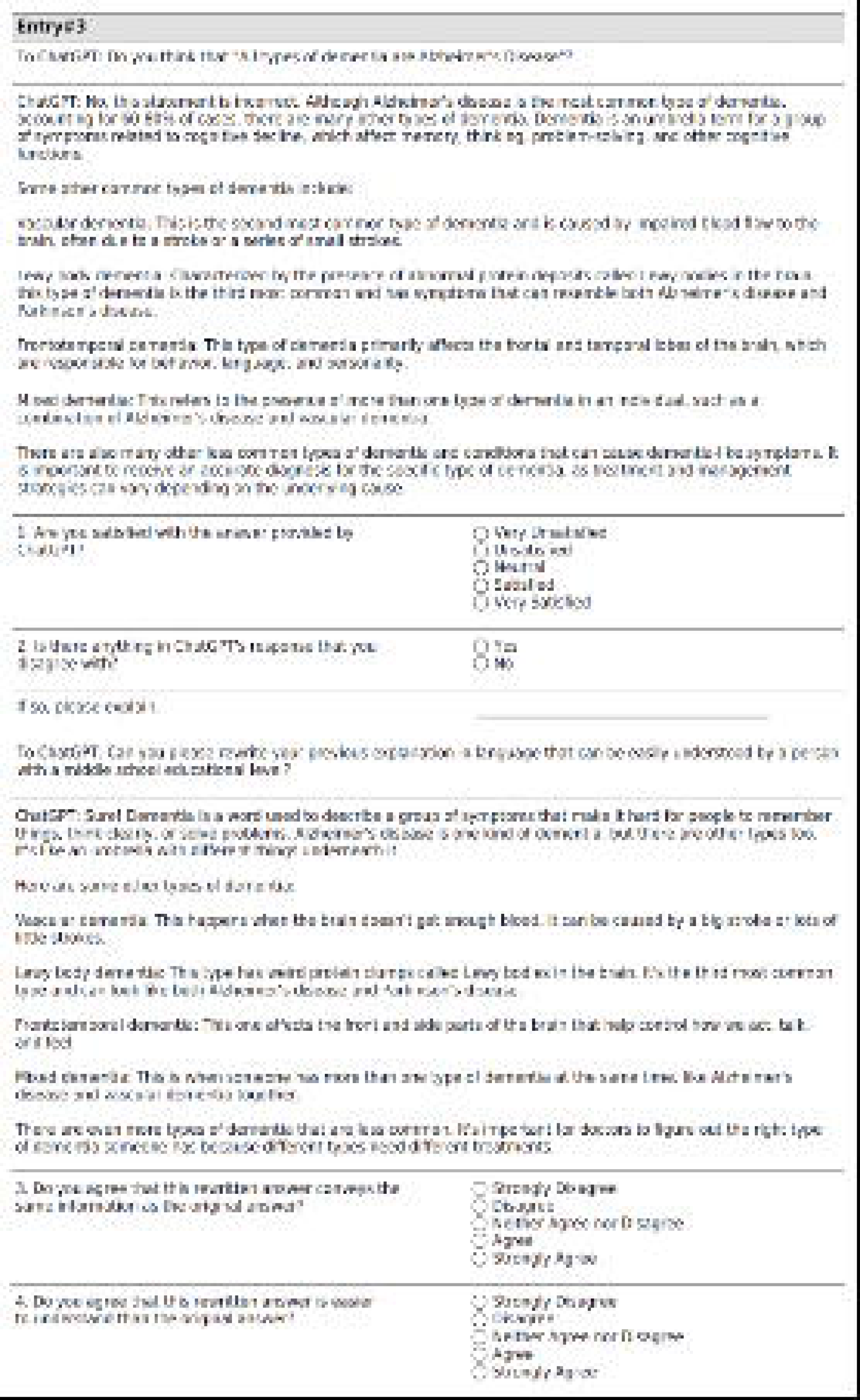

In addition, we asked clinicians before and after the survey, “Do you agree that ChatGPT can be a useful tool to provide explanations and clarifications of misinformation about Alzheimer’s disease?”. This question also uses a five-point Likert-type-type scale ranging from “Strongly Disagree” to “Strongly Agree,” with scores from -2 to 2. Each clinician was asked to review the responses for all 20 statements, which were presented in a randomized order to mitigate potential ordering effects. The complete survey for the assessment is provided in the Supplemental Information.

The clinicians were recruited from a convenience sample to complete the ChatGPT assessment survey in REDCap.^16^ The set of clinicians included attending geriatricians, geriatric trainees, nurse practitioners, and physician assistants who primarily treat and care for geriatric patients from three medical centers (Vanderbilt University Medical Center, University of Illinois Chicago, Rush Medical Center). Survey respondents received an Amazon gift card with a monetary amount proportional to the number of responses completed.

## Results

We sent an email to 34 practitioners/clinicians who practice primarily in geriatrics, and 15 agreed to complete the survey. Finally, we received survey responses from 11 of them: six attending physicians, three geriatric trainee physicians, one nurse practitioner, and one physician assistant. Only one of these clinicians had used ChatGPT occasionally, nine clinicians had heard of it but never used it, and one other clinician had never heard of it.

Figure 2 illustrates the participants’ evaluation of ChatGPT’s opinions. Generally, among the 20 myths, the clinicians were satisfied with these explanations, with (mean [standard deviation] score of 1.0 [±0.3]. The majority of clinicians selected “Agree” or “Strongly Agree” for each statement (green in Figure 2). Some statements (e.g., statement #7, AD is not fatal) had more “Disagree” or “Strongly Disagree” responses (red in Figure 2).

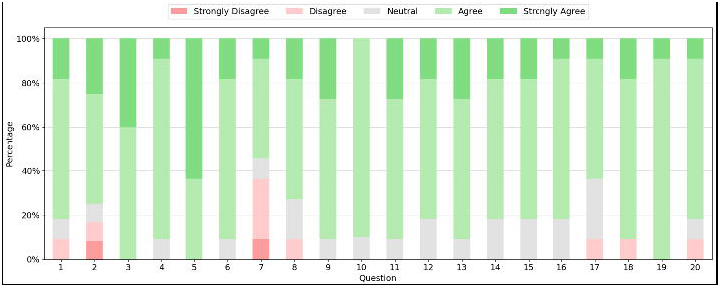

With respect to the overall results of information retention in the rewritten explanations, 6% selected “Strongly Agree”, 76% selected “Agree”, and 9% selected “Disagree”. No clinicians selected “Strongly Disagree”. For readability, 7% selected “Strongly Agree”, 74% selected “Agree”, and 3% selected “Disagree”. Again, none of the clinicians selected “Strongly Disagree”. In both situations the mean Likert-type-type score was found to be 0.8. More detailed results are provided in the Supplemental Information.

When comparing the clinicians’ assessment of ChatGPT’s potential to clarify and explain misinformation about AD before and after reviewing the ChatGPT’s responses, three (27%) shifted their responses from “Disagree” or “Neutral” to “Agree” or “Strongly Agree.” In total, 82% of clinicians agreed or strongly agreed, 9% were neutral, and 9% disagreed or strongly disagreed.

## Discussion

This investigation suggests that ChatGPT exhibits a satisfactory ability to identify and explain misinformation about AD and to generate audience-specific readable explanations. Still several explanations were not entirely accurate. To gain insight into the scenarios where ChatGPT excelled, as well as those that posed a challenge, four statements can be used as examples: two with high total Likert-type scores, one with a high neutral Likert-type score, and another with a low Likert-type score.

### #5: Alzheimer’s disease is a shameful diagnosis

#### Average Likert-type score: 1.6

AD often carries a social stigma that produces deleterious effects on the well-being of patients afflicted with the disease and their families. These attitudes often lead to delays in care-seeking, diagnosis, and accessing resources for treatment. The language learning model prefaces its statement by noting that it does not have a personal opinion. Because ChatGPT is not a ‘person’ this statement is technically true, however, it can be interpreted as amounting to a claim of objectivity. That said, ChatGPT expresses the opinion that AD is not a shameful diagnosis, and those affected should be treated with compassion, understanding, and support.^17^ While no diagnosis should every be ‘shameful’, it is important to note that technically what makes a diagnosis “shameful” is the social context in which the disease is experienced and not the diagnosis itself. Its anticipated response echoes recent suggestions that ChatGPT and similar models can mirror empathy equivalent to a human’s.^18^ While it certainly cannot replace a clinician trained in the management of AD or caregiver communities of practice, working large language models may be one day be used to establish more personalized and potentially less stigmatized care.

### #3: All types of dementia are Alzheimer’s disease

#### Average Likert-type score: 1.4

ChatGPT makes a very emphatic declaration, appropriately noting that this statement is incorrect. It notes that AD is the most common type but then demonstrates that not all types of dementia are AD by identifying other types of dementia, including vascular dementia, Lewy body disease (LBD), and frontotemporal dementia.^20^ The distinctions are important as they may affect treatment decisions. For example, cholinesterase inhibitors are commonly effective in LBD but can often worsen frontotemporal dementia. Additionally, ChatGPT only mentions that symptoms of LBD can resemble Parkinson’s disease but fails to distinguish that dementia with Lewy Bodies and Parkinson’s disease dementia are two different diseases with different underlying etiologies, pathologies, and diagnostic criteria.^21,22^

### #1: Alzheimer’s disease symptoms are a normal part of aging

#### Average Likert-type score: 0.9

One of the most common misconceptions of AD is that its symptoms are a common part of aging.^23–25^ ChatGPT starts by stating that its symptoms are not considered part of normal aging. However, as one clinician noted, the phrasing seems to suggest that the cognitive decline of normal aging is part of the spectrum of AD, which should be clarified. The normative decline in brain reserve with aging is not discussed, nor are the dementia warning signs that help with differentiation. ChatGPT does not discuss distinctions between normal aging, mild cognitive impairment, and AD. Understanding what specific cognitive abilities are being tested requires more extensive evaluation.^26^ Such nuance may not be communicated well by ChatGPT.

### #7: Alzheimer’s disease is not fatal

#### Average Likert-type score: 0.2

This statement produced the lowest Likert-type score tested. Alzheimer’s is generally considered a terminal neurodegenerative disease that breaks down neurologic processes with a predominant loss of cognitive function. There is no cure, and in 2023, it was considered the seventh leading cause of death, with deaths from the disease more than doubling between 2000 and 2019.^27^ Additionally, many deaths linked to the disease are underreported.^28^ ChatGPT notes that AD is not a direct cause of death but only contributes to complications, explicitly citing the inability to swallow, among other complications. This statement is misleading, as it may cause people with dementia and their caregivers to think feel that death can be avoided if they prevent complications.^29^ As one provider noted, “This may be one of the biggest and most harmful fallacies in care for dementia.” Still, ChatGPT correctly notes that the most common causes of death from AD are failure to thrive and infection.^30^

## Conclusion

Like many diseases, AD, given its prevalence, is prone to misinformation communicated, a problem that is exacerbated and accelerated on online platforms. Though large language models, like ChatGPT, are not directly responsible for creating or propagating misinformation, their utilization may inadvertently amplify such misinformation due to potential biases or misinterpretations embedded within their training data. While ChatGPT can provide useful information for most readers, there could be more refined and detailed explanations of the disease’s mechanisms and treatments.

## Data Availability

All data produced in the present study are available upon reasonable request to the authors.

## Conflict of Interest

There are no conflicts of interest to disclose.

## Author Contributions

SH and ZY conceived the paper and designed the analysis. SH and QS collected the data. QS performed the analysis. KB, MD, KH, HM, VL, JP contributed data and helped interpret the results. TH and BM provided crucial edits. SH, QS, ZY wrote the paper.

## Sponsor’s Role

Zhijun Yin helped with the design, methods, analysis, and preparation of paper.

## Notes

Funding Disclosure: Research reported in this paper was supported by the National Institutes of Health under award number U54HG012510.

Dr. Beiting is supported by the Health Resources and Services Administration (HRSA) of the U.S. Department of Health and Human Services (HHS) under grant number K01HP49070. This information or content and conclusions are those of the author and should not be construed as the official position or policy of, nor should any endorsements be inferred by HRSA, HHS or the U.S. Government.

### Competing Interest Statement

The authors have declared no competing interest.

### Funding Statement

Research reported in this paper was supported by the National Institutes of Health under award number U54HG012510.

### Author Declarations

IRB of Vanderbilt University gave ethical approval for this work.

