## Supplemental Information for "Fact Check: Assessing the Response of ChatGPT to Alzheimer’s Disease Statements with Varying Degrees of Misinformation"

This document provides supplemental evidence to the research findings of the main manuscript. This document is split into two sections. The first section presents the characteristics of the Amazon Mechanical Turk (AMT) participants, the degree to which they agreed with the 20 statements about Alzheimer's Disease (AD), and the regression analysis on the association between participants' characteristics and their agreements with the statements. The second component includes clinicians' assessment of ChatGPT's rewritten explanations and the full ChatGPT's responses to 20 statements.

#### A. AMT participants opinion of the 20 AD statements

We selected 20 commonly held myths related to Alzheimer's Disease and validated them using a survey distributed to 200 participants via Amazon Mechanical Turk. AMT is an online platform that outsources various tasks, including surveys, to a distributed workforce. Each participant was awarded $1 for their contribution.

The survey utilized a 5-point Likert scale for assessing the participants' agreement or disagreement with the presented myths. Here, the options "Strongly Disagree" and "Disagree" were assigned value of 1, "Neutral" was assigned value of 0, while "Agree" and "Strongly Agree" were assigned value of -1. Given that all 20 myths were formulated with certain degrees of misinformation, the correct responses should be either “Strongly Disagree” or “Disagree”. Therefore, a positive Likert score indicates a good understanding of the myths. This approach allows us to validate the likelihood of these 20 myths being accepted as common misconceptions about AD within a broad community.

In addition to the assessment of these myths, the survey also gathered personal information about the participants, such as their age, gender, race, ethnicity, hearing about AD, whether they have cared for AD patients, and whether they received payment for their caregiving services. Subsequently, a linear regression analysis was conducted to elucidate the potential relationships between the participants' personal information and their level of agreement, in terms of the average Likert score, with the myths.

#### Characteristics of AMT participants

**Table S-1** Characteristic table for 200 participants for AMT survey

| Characteristic | Total n=200 |
|---|---|
| Age in years, mean (range) | 34.8 (22 - 68) |
| Gender, n (%) | |
| Male | 139 (69.5) |
| Female | 60 (30.0) |

| | |
|---|---|
| Undifferentiated | 1 (0.5) |
| Race, n (%) | |
| White | 179 (89.5) |
| Black or African American | 8 (4.0) |
| American Indian or Alaska Native | 7 (3.5) |
| Asian | 5 (2.5) |
| Native Hawaiian or Other Pacific Islander | 1 (0.5) |
| Ethnicity, n (%) | |
| Not Hispanic or Latino | 172 (86.0) |
| Hispanic or Latino | 28 (14.0) |
| Education, n (%) | |
| Some high school but did not graduate | 4 (2.0) |
| High school or GED | 16 (8.0) |
| Some college or 2-year degree | 22 (11.0) |
| 4-year college graduate | 140 (70.0) |
| More than 4-year college degree | 18 (9.0) |
| Hearing about AD, n (%) | |
| Yes | 169 (84.5) |
| No | 31 (15.5) |
| Caring for an AD patient, n (%) | |
| Yes | 124 (62.0) |
| No | 45 (22.5) |
| Do not apply | 31 (15.5) |
| Caring for an AD patient with payment, n (%) | |
| Yes | 89 (44.5) |
| No | 35 (17.5) |
| Do not apply | 76 (38.0) |

**Table S-1** summarizes the characteristics of the 200 AMT participants. The 200 AMT participants were mostly male (69.5%), identified as white (89.5%), not Hispanic or Latino ethnicity (86.0%), and held a 4-year college degree (70%). There were 169(85%) participants who reported hearing about AD and 124 of them had been involved in caring for a person with AD. Among the caregivers, 89 participants indicated that they received payment for providing the caregiving service. It should be noted that we designed the questions regarding "Hearing about Alzheimer's Disease" (denoted by "Hearing about AD"), "Caring for an AD patient" (denoted by "Caregiver"), and "Caring for an AD patient with payment" (denoted by "Caregiver with Payment") with a nested relationship. Participants who responded with 'No' to a preceding question were not able to respond to the subsequent one. For completeness, these participants are included in the "Do not apply" category in Table S-1.

### Regression model parameters

We performed a regression analysis to examine which characteristics of AMT participants are statistically significantly associated with their agreement of statements at a significance level of 0.05. We used the average score, derived from the five-point Likert scale of agreement for the 20 AD statements for each participant, as the dependent variable in the model. We performed a binary encoding on categorical predictors. Specifically, we encoded the dominant category of each categorical predictor as 1 and the remaining categories as 0. For example, for race, we represent the 'White' category as 1 and all the other categories by 0. Since "Hearing about AD", "Caregiver", and "Caregiver with Payment" are highly correlated with each other, we incorporated them into three linear regression models, $M_H$, $M_C$, $M_P$, separately.

### Regression model results

**Table S-2** Results for the three regression models. The asterisk (*) indicates that the corresponding predictor is significantly associated with the average Likert score.

| | $M_H$ | | $M_C$ | | $M_P$ | |
|---|---|---|---|---|---|---|
| **Predictors** | **Coeff.** | **Pr(>\|t\|)** | **Coeff.** | **Pr(>\|t\|)** | **Coeff.** | **Pr(>\|t\|)** |
| Age | -6.13 | 0.13 | -5.36 | 0.16 | -3.78 | 0.31 |
| Gender - Male | -2.36 | 0.14 | -2.14 | 0.16 | -2.26 | 0.13 |
| Race - White | 1.86 | 0.45 | 0.98 | 0.68 | 0.22 | 0.92 |
| Ethnicity - Not Hispanic or Latino | 1.99 | 0.36 | 1.12 | 0.59 | 1.48 | 0.46 |
| Education - 4-year college graduate or more | 0.06 | 0.06 | 3.16 | 0.07 | 2.52 | 0.14 |
| Hearing about AD | -1.70 | 0.40 | | | | |
| **Caregiver** | | | **6.49** | **<0.001 *** | | |
| **Caregiver with Payment** | | | | | **7.96** | **<0.001 *** |
| (Intercept) | 3.75 | 0.31 | -0.93 | 0.77 | 0.33 | 0.91 |

**Table S-2** presents the outcomes of the linear regression analyses. It was found that the variables "Caregiver" and "Caregiver with Payment" have a significant relationship with the outcome ($p < 0.001$). The coefficients of these variables were both positive, with the value of 6.49 and 7.96, respectively. This suggests that misinformation about AD was less prevalent among those with caregiving experience for AD patients, particularly for those received payment for their caregiving services. It is possible that individuals who have never cared for an Alzheimer's patient might lack first-hand experience and, therefore, might be more susceptible to misinformation.

### Summary figure of the survey response

***Figure S-1*** illustrates the proportion of the Likert scale responses for each of 20 Alzheimer's Disease misinformation statements. The x-axis depicts the percentage of participants' selection, while the y-axis enumerates the statements. Notably, the most frequently chosen responses for each myth among the AMT participants were "Agree" and "Neutral", representing a mean(SD) of 33%(±4%) and 32%(±4%) respectively. This suggests that the 20 pre-selected misconceptions of AD were not thoroughly understood among the participant population. Only 36% of the participants correctly responded to all statements, with an average Likert scale score of -0.2 (±0.1) for all statements, indicating these 20 statements are accepted as common misconceptions. Specifically, the highest average Likert score (0.04) is seen for statement #9 (AD patients cannot get depressed) and #18 (Vaccines increase the risk of AD). Conversely, the lowest average Likert score (-0.33) is noticed for statement #14 (Prescription medications can reverse the progression to AD).

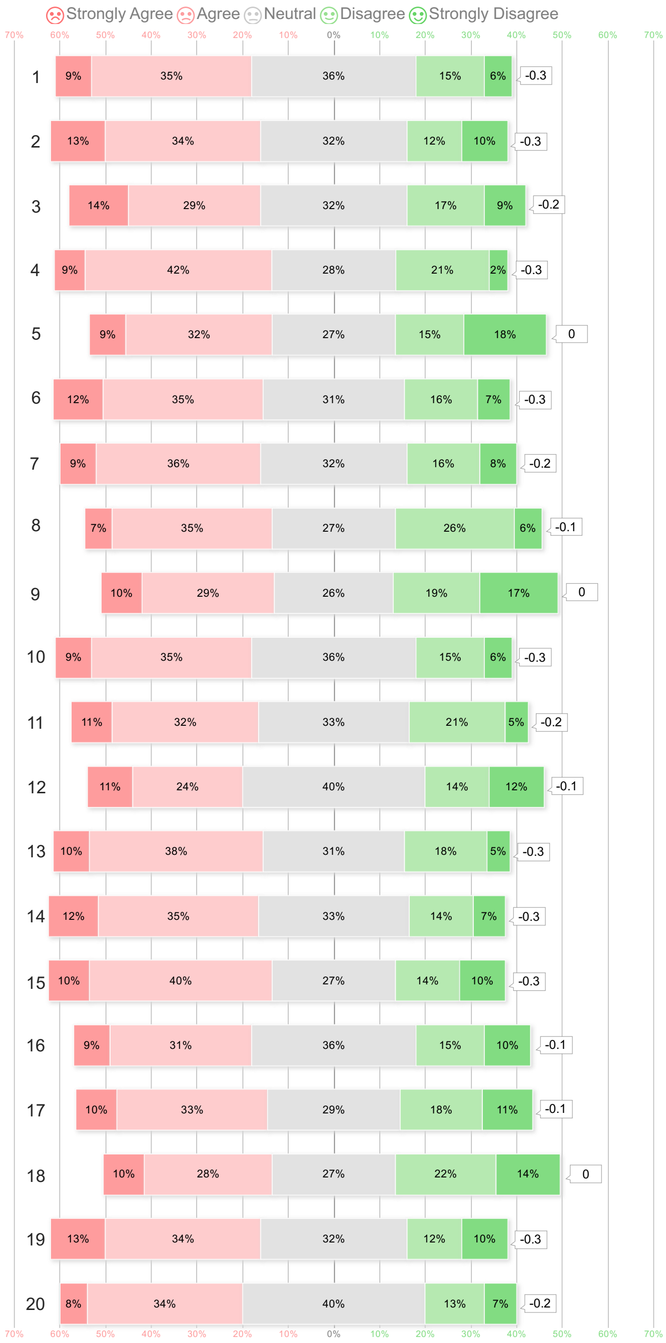


**Figure S-1** AMT participants survey responses for agreement of 20 myths. Strongly Disagree and Disagreed responses are shaded in red while Agree and Strongly Agreed responses are shaded in green.

### B. Clinician's Assessment of ChatGPT's explanations

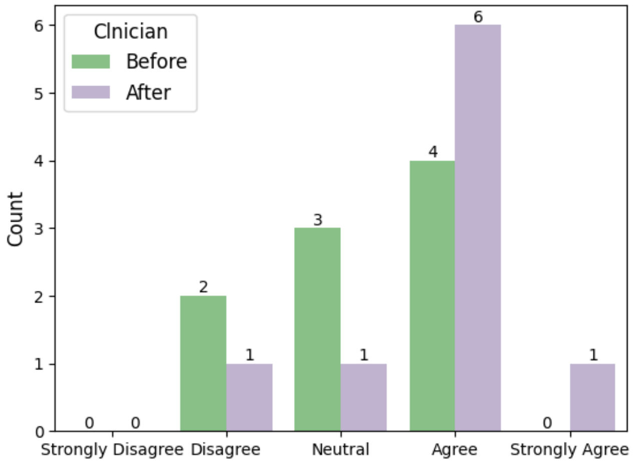


**Figure S-2** Distribution of responses to the question "Do you agree that ChatGPT can be a useful tool to provide explanations and clarifications of misinformation about Alzheimer's disease?" before and after reviewing the ChatGPT explanations.

After reviewing ChatGPT's responses, the clinicians indicated an overall positive attitude, corroborated by the reasons they specified in their comment. Within their comments, the clarity and conciseness of the information provided were consistently emphasized. However, they also pointed out several areas for potential improvement. These included a need for the communication of more nuanced details, as well as the provision of personalized recommendations according to the specific circumstances. A minority of the feedback indicated the presence of incomplete or potentially misleading information. The clinicians reiterated the importance for detailed proofreading and professional healthcare counseling to address any concerns. However, this did not undermine the clinicians' overall approval of the ChatGPT as a useful tool, and they expressed an expectation of continuous improvement in its efficiency.

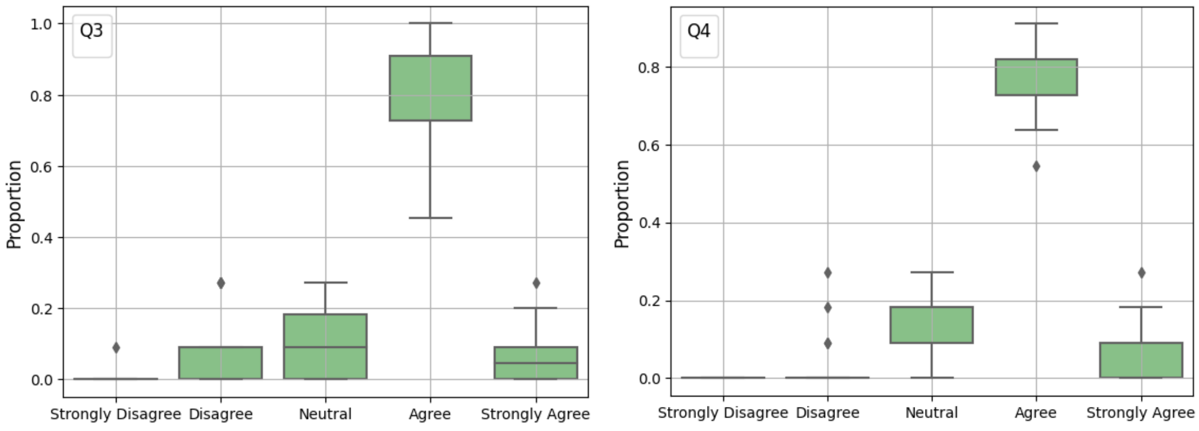


**Figure S-3** (Left): Distribution of responses to the question "Do you agree that this rewritten explanation conveys the same information as the original answer?"; (Right): Choice distribution of the question "Do you agree that this rewritten explanation is easier to understand than the original answer?"

**Figures S-3** illustrates the distribution of responses from clinicians for rewritten explanations of 20 statements: the extent of information retained from the ChatGPT's rewritten explanation, and the readability of the rewritten explanation when compared with the original one. It is evident that the mean proportion of participants selecting "Agree" consistently exceeds 0.5**,** signifying a general positive trend in the evaluation of ChatGPT's rewritten explanations.

ChatGPT demonstrated an ability to generate responses that meet diverse levels of background knowledge. As noted by one clinician, “[the] rewritten explanation is too cursive but usually seemed to encompass gist of information”. This suggests a potential for ChatGPT to generate accurate information across a wide range of educational backgrounds, with competence in distilling complex medical concepts into digestible information.

### C. Full Responses to 20 Myths from ChatGPT

#### Myth #1

**To ChatGPT:** Do you think that "Alzheimer's Disease symptoms are a normal part of aging"?

**ChatGPT:** As an AI language model, I don't have personal opinions, but I can provide you with information based on scientific consensus. It is not accurate to say that Alzheimer's Disease symptoms are a normal part of aging. While it is true that aging is the most significant risk factor for Alzheimer's Disease, the symptoms and cognitive decline associated with it are not considered part of normal aging. Alzheimer's Disease is a progressive neurological disorder that leads to the degeneration and death of brain cells. It is the most common cause of dementia, a term used to describe a group of symptoms that affect cognitive function, such as memory loss, difficulty thinking, problem-solving, and language skills. Although mild forgetfulness and cognitive changes can be a part of normal aging, the symptoms of Alzheimer's Disease are much more severe and debilitating. It is important to differentiate between normal age-related memory decline and the more serious cognitive decline associated with Alzheimer's Disease. If you or someone you know is experiencing concerning memory problems or other cognitive issues, it is crucial to consult a healthcare professional for a proper evaluation and diagnosis. Early detection and intervention can help manage the symptoms and potentially slow the progression of the disease.

**To ChatGPT:** Can you please rewrite your previous explanation in language that can be easily understood by a person with a middle school educational level?

**ChatGPT:** Sure! Alzheimer's Disease symptoms are not a normal part of getting older. Even though people have a higher chance of getting Alzheimer's as they age, the problems it causes are not just because of getting older. Alzheimer's Disease is a brain problem that gets worse over time. It makes it hard for people to remember things, think clearly, and communicate. This is different from the small memory issues that can happen as we grow older. Alzheimer's Disease causes much bigger problems than what usually happens when people age. If someone is having a lot of trouble with their memory or thinking, they should talk to a doctor. The doctor can help figure out what is going on and find ways to help them.

### Myth #2

**To ChatGPT:** Do you think that "Symptoms of memory loss must be Alzheimer's Disease"?

**ChatGPT:** As an AI language model, I don't have personal opinions or beliefs. However, I can provide you with some information on the topic. It is incorrect to assume that symptoms of memory loss must be Alzheimer's disease. Memory loss can be caused by various factors, and Alzheimer's is just one of the potential causes. Other possible causes of memory loss include:

- Age-related cognitive decline: As people age, they may experience mild cognitive decline, which can lead to memory loss. This is a normal part of aging and not necessarily indicative of Alzheimer's or another form of dementia.
- Stress and anxiety: High levels of stress and anxiety can impair memory function temporarily.
- Depression: Depression can cause cognitive issues, including memory problems.
- Sleep disorders: Poor sleep quality or sleep disorders, such as sleep apnea, can lead to memory loss.
- Medications: Certain medications can have side effects that affect memory.
- Vitamin deficiencies: Deficiencies in certain vitamins, such as vitamin B12, can lead to memory problems.
- Other medical conditions: Conditions like thyroid problems, infections, or brain tumors can cause memory loss.
- Other forms of dementia: Alzheimer's disease is just one type of dementia. There are other forms, such as vascular dementia, Lewy body dementia, and frontotemporal dementia, which can also cause memory loss.

It is essential to consult a healthcare professional if you or someone you know is experiencing memory loss. Proper evaluation and diagnosis can help determine the underlying cause and appropriate treatment.

**To ChatGPT:** Can you please rewrite your previous explanation in language that can be easily understood by a person with a middle school educational level?

**ChatGPT:** Of course! Memory loss doesn't always mean someone has Alzheimer's disease. There are many reasons someone might have trouble remembering things. Here are a few examples:

- Getting older: As people grow older, it's normal for their memory to get a little worse. This doesn't always mean they have Alzheimer's.
- Stress and worry: When someone is really stressed or worried, they might have a hard time remembering things.
- Feeling sad: Sometimes when people are very sad or depressed, their memory can be affected.
- Not sleeping well: If someone doesn't sleep well or has a problem with their sleep, it can make their memory worse.
- Medicines: Some medicines can make it harder to remember things.

- Not enough vitamins: If someone doesn't have enough of certain vitamins, like vitamin B12, their memory might not be as good.
- Health problems: Some health issues, like problems with the thyroid or even a brain tumor, can cause memory loss.
- Different kinds of dementia: Alzheimer's is just one type of dementia. There are others, like vascular dementia or Lewy body dementia, that can also cause memory problems.

If you or someone you know is having trouble with memory, it's important to talk to a doctor. They can help figure out what's causing the memory loss and how to treat it.

#### Myth #3

**To ChatGPT:** Do you think that "All types of dementia are Alzheimer's Disease"?

**ChatGPT:** No, this statement is incorrect. Although Alzheimer's disease is the most common type of dementia, accounting for 60-80% of cases, there are many other types of dementia. Dementia is an umbrella term for a group of symptoms related to cognitive decline, which affect memory, thinking, problem-solving, and other cognitive functions. Some other common types of dementia include:

- Vascular dementia: This is the second most common type of dementia and is caused by impaired blood flow to the brain, often due to a stroke or a series of small strokes.
- Lewy body dementia: Characterized by the presence of abnormal protein deposits called Lewy bodies in the brain, this type of dementia is the third most common and has symptoms that can resemble both Alzheimer's disease and Parkinson's disease.
- Frontotemporal dementia: This type of dementia primarily affects the frontal and temporal lobes of the brain, which are responsible for behavior, language, and personality.
- Mixed dementia: This refers to the presence of more than one type of dementia in an individual, such as a combination of Alzheimer's disease and vascular dementia.

There are also many other less common types of dementia and conditions that can cause dementia-like symptoms. It is important to receive an accurate diagnosis for the specific type of dementia, as treatment and management strategies can vary depending on the underlying cause.

**To ChatGPT:** Can you please rewrite your previous explanation in language that can be easily understood by a person with a middle school educational level?

**ChatGPT:** Sure! Dementia is a word used to describe a group of symptoms that make it hard for people to remember things, think clearly, or solve problems. Alzheimer's disease is one kind of dementia, but there are other types too. It's like an umbrella with different things underneath it. Here are some other types of dementia:

- Vascular dementia: This happens when the brain doesn't get enough blood. It can be caused by a big stroke or lots of little strokes.
- Lewy body dementia: This type has weird protein clumps called Lewy bodies in the brain. It’s the third most common type and can look like both Alzheimer’s disease and Parkinson’s disease.

- Frontotemporal dementia: This one affects the front and side parts of the brain that help control how we act, talk, and feel.
- Mixed dementia: This is when someone has more than one type of dementia at the same time, like Alzheimer's disease and vascular dementia together.

There are even more types of dementia that are less common. It's important for doctors to figure out the right type of dementia someone has because different types need different treatments.

#### Myth #4

**To ChatGPT:** Do you think that "All Alzheimer's Disease patients are disoriented, look confused, and act outside social norms"?

**ChatGPT:** As an AI language model, I don't possess personal opinions, but I can provide information based on existing knowledge. The statement "All Alzheimer's disease patients are disoriented, look confused, act outside social norms" is an overgeneralization. Although many Alzheimer's patients may exhibit these symptoms, it's important to remember that the disease presents itself differently in each individual. Alzheimer's disease is a progressive neurodegenerative disorder that affects memory, cognitive function, and behavior. In the early stages, symptoms may be mild and not noticeable to others. As the disease progresses, symptoms become more pronounced and can include disorientation, confusion, and changes in behavior. However, the severity and progression of these symptoms can vary significantly between individuals. It is also crucial to note that Alzheimer's disease is not the only cause of cognitive decline or disorientation in [older] individuals. Other factors, such as medications, other health conditions, and age-related changes, can also contribute to these symptoms. Therefore, it is important to approach this topic with sensitivity and avoid making sweeping generalizations.

**To ChatGPT:** Can you please rewrite your previous explanation in language that can be easily understood by a person with a middle school educational level?

**ChatGPT:** Sure! Alzheimer's disease is a brain illness that makes it hard for people to remember things, think clearly, and act normally. But not everyone with Alzheimer's shows the same signs. Some people might be confused or act strangely, while others might not show these signs as much. It's important to remember that Alzheimer's is different for everyone. Sometimes, people can have trouble with their memory or thinking because of other reasons, like taking certain medicines or having other health problems. So, saying that all people with Alzheimer's are confused or act strangely isn't true for everyone. We should be careful not to assume that everyone with Alzheimer's is the same.

#### Myth #5

**To ChatGPT:** Do you think that "Alzheimer's Disease is a shameful diagnosis"?

**ChatGPT:** As an AI language model, I don't have personal opinions or feelings. However, I can share that there is no reason to consider Alzheimer's disease or any other medical condition as a "shameful diagnosis." Alzheimer's disease is a neurodegenerative disorder that affects memory, thinking, and behavior. It is a medical condition that should be treated with compassion, understanding, and support. Stigmatizing Alzheimer's or any other medical condition can have harmful effects on individuals with the disease and their families, as it may lead to isolation, delay in seeking help, and diminished quality of life. It is essential to raise awareness about the disease, promote understanding, and support research to improve treatment options and quality of life for those affected by Alzheimer's disease.

**To ChatGPT:** Can you please rewrite your previous explanation in language that can be easily understood by a person with a middle school educational level?

**ChatGPT:** Sure! Alzheimer's disease is a sickness that affects a person's memory, thinking, and behavior. There is no reason to feel ashamed about it or any other health problem. We should treat people with Alzheimer's with kindness, care, and support. Saying that Alzheimer's is shameful can hurt people with the disease and their families. It can make them feel alone and stop them from asking for help. We need to teach people about Alzheimer's and help find better ways to treat it and take care of those who have it.

### Myth #6

**To ChatGPT:** Do you think that "An at-home genetic test can tell me if I have (or will have) Alzheimer's Disease"?

**ChatGPT:** At-home genetic tests can provide some information about your risk of developing Alzheimer's disease, but they cannot definitively tell you if you have or will develop the condition. Alzheimer's disease is a complex neurodegenerative disorder with both genetic and environmental factors contributing to its development. While certain genes have been associated with an increased risk of Alzheimer's, having these genetic markers does not guarantee that you will develop the disease. One well-known genetic risk factor for Alzheimer's is the APOE gene. There are three common versions of this gene: APOE ε2, APOE ε3, and APOE ε4. Having one or two copies of the APOE ε4 variant is associated with a higher risk of developing Alzheimer's, while having the APOE ε2 variant is associated with a lower risk. However, even if you have an APOE ε4 variant, it does not mean you will definitely develop Alzheimer's. Many people with the APOE ε4 variant never develop the disease, and some people without the variant still do. At-home genetic tests can determine which variants of the APOE gene you have, but interpreting the results is not straightforward. Genetic testing for Alzheimer's risk is only one piece of the puzzle, and other factors such as age, family history, lifestyle, and overall health also play a role in determining your risk. Moreover, genetic testing cannot predict when or how severe the disease may be if it develops. If you are concerned about your risk of developing Alzheimer's disease, it is important to discuss your concerns with a healthcare professional, who can provide more accurate and personalized advice based on your medical history and other factors.

**To ChatGPT:** Can you please rewrite your previous explanation in language that can be easily understood by a person with a middle school educational level?

**ChatGPT:** At-home genetic tests can give you some information about your chances of getting Alzheimer's disease, but they can't tell you for sure if you have it or will get it. Alzheimer's is a brain disease that can be caused by a mix of genes and things that happen in your life, like your environment. Some genes can make you more likely to get Alzheimer's, but having these genes doesn't mean you'll definitely get the disease. There is a gene called APOE that is linked to Alzheimer's. There are three common types of this gene: APOE ε2, APOE ε3, and APOE ε4. If you have the APOE ε4 type, you have a higher chance of getting Alzheimer's. If you have the APOE ε2 type, your chances are lower. But even if you have the APOE ε4 type, you might not get Alzheimer's, and if you don't have it, you still could get the disease. At-home genetic tests can tell you which type of APOE gene you have, but the results can be hard to understand. There are many other things, like age, family history, how you live, and your overall health, that can also affect your chances of getting Alzheimer's. Genetic tests can't tell you when or how bad the disease might be if you do get it. If you're worried about your chances of getting Alzheimer's, it's important to talk to a doctor. They can give you more specific advice based on your personal situation.

#### Myth #7

**To ChatGPT:** Do you think that "Alzheimer's Disease is not fatal"?

**ChatGPT:** As an AI language model, I don't have personal opinions, but I can provide you with information on the topic. Alzheimer's disease is a progressive neurodegenerative disorder that affects a person's memory, thinking, and behavior. While the disease itself is not the direct cause of death, it does have a significant impact on a person's health and can lead to complications that may ultimately be fatal. Individuals with Alzheimer's often experience a decline in cognitive and physical function, which can make it difficult for them to perform daily tasks and self-care. As the disease progresses, patients become more susceptible to infections, falls, and other health issues. In advanced stages, Alzheimer's patients may have difficulty swallowing, which can lead to malnutrition, dehydration, and aspiration pneumonia - a condition where food or liquid enters the lungs, leading to infection. In summary, while Alzheimer's disease itself may not be directly fatal, it contributes to a decline in overall health and can lead to complications that can be life-threatening.

**To ChatGPT:** Can you please rewrite your previous explanation in language that can be easily understood by a person with a middle school educational level?

**ChatGPT:** Alzheimer's disease is a brain sickness that makes it hard for people to remember things, think clearly, and take care of themselves. Even though Alzheimer's doesn't cause death directly, it can lead to other health problems that can be very dangerous. As Alzheimer's gets worse, people have a harder time doing everyday tasks and staying healthy. They can get sick more easily, fall down, or have other accidents. When the disease is really bad, people might have trouble eating and drinking, which can make them weak and lead to more sickness. In short, Alzheimer's disease doesn't directly cause death, but it can make a person's health worse and lead to other dangerous problems that could be life-threatening.

#### Myth #8

**To ChatGPT:** Do you think that "Alzheimer's Disease patients are close to death"?

**ChatGPT:** As an AI language model, I don't have personal opinions or beliefs. However, I can provide you with information on Alzheimer's disease and its progression. Alzheimer's disease is a progressive neurological disorder that affects memory, thinking, and behavior. The progression of Alzheimer's varies from person to person, and the life expectancy of individuals with the disease can also differ significantly. In the early stages of Alzheimer's, a person might experience mild memory loss and cognitive decline, but they can still function independently. As the disease progresses, symptoms become more severe, and individuals may require assistance with daily activities. In the later stages of Alzheimer's, a person may be unable to communicate, recognize loved ones, or take care of themselves. It's important to note that Alzheimer's disease itself is not usually the direct cause of death; rather, complications associated with the disease, such as infections or pneumonia, are more likely to be fatal. Life expectancy after an Alzheimer's diagnosis can range from 3 to 20 years, depending on various factors such as age, general health, and the severity of symptoms. It is not accurate to say that all Alzheimer's patients are close to death, as the disease can progress at different rates for each individual. If you have concerns about a loved one with Alzheimer's disease or need more information about the condition, it's crucial to consult with a healthcare professional for accurate guidance and support.

**To ChatGPT:** Can you please rewrite your previous explanation in language that can be easily understood by a person with a middle school educational level?

**ChatGPT:** Sure, I can simplify the explanation for you. Alzheimer's disease is a brain problem that makes it hard for people to remember things, think clearly, and behave normally. The disease affects different people in different ways, and how long they live after being diagnosed can also be different. When someone first gets Alzheimer's, they might forget things sometimes, but they can still do most things on their own. As the disease gets worse, they will have more trouble thinking and doing everyday tasks. In the last stages of Alzheimer's, a person may not be able to talk, know who their family and friends are, or take care of themselves. But it's not usually the Alzheimer's itself that causes people to die. Instead, they often get sick with other problems, like infections or lung problems, because of the Alzheimer's. After being diagnosed with Alzheimer's, a person might live anywhere from 3 to 20 years. This depends on things like their age, overall health, and how bad their Alzheimer's symptoms are. So, it's not right to say that all people with Alzheimer's are close to dying, because the disease can get worse at different speeds for different people. If you're worried about someone with Alzheimer's or want to learn more, it's important to talk to a doctor or another expert who can give you the right information and help.

#### Myth #9

**To ChatGPT:** Do you think that "Alzheimer's Disease patients cannot get depressed"?

**ChatGPT:** As an AI language model, I don't have personal opinions or beliefs, but I can provide you with information based on existing knowledge. It is not accurate to say that Alzheimer's disease patients cannot get depressed. In fact, individuals with Alzheimer's disease can and do experience depression. Depression is a common neuropsychiatric symptom of Alzheimer's disease, affecting approximately 30-50% of patients at some point during their illness. Depression in Alzheimer's patients can manifest as persistent sadness, social withdrawal, loss of interest in activities they once enjoyed, irritability, and changes in sleep and appetite patterns. It can be challenging to identify depression in individuals with Alzheimer's, as some of the symptoms overlap with those of dementia. It is essential to recognize and treat depression in Alzheimer's patients, as it can exacerbate cognitive decline, decrease quality of life, and increase caregiver burden. Treatment may include medication, therapy, or a combination of both, depending on the severity of the depression and the patient's overall health.

**To ChatGPT:** Can you please rewrite your previous explanation in language that can be easily understood by a person with a middle school educational level?

**ChatGPT:** Sure! People with Alzheimer's disease can definitely feel sad or depressed. In fact, many people with Alzheimer's go through periods of feeling very sad or not enjoying things they used to like. Sometimes, it's hard to tell if someone with Alzheimer's is feeling depressed because the signs can be similar to the memory problems they already have. But it's important to know if someone is feeling this way, because being sad can make their memory problems worse and make it harder for them to enjoy life. To help someone with Alzheimer's who is feeling depressed, doctors might give them medicine, talk with them about their feelings, or both. This can help them feel better and improve their life.

### Myth #10

**To ChatGPT:** Do you think that "I will develop Alzheimer's Disease if my parent has it"?

**ChatGPT:** I am not a doctor, but I can provide you with some general information about Alzheimer's disease and genetic risk factors. Having a parent with Alzheimer's disease does not necessarily mean that you will develop the disease. While genetics can play a role in the development of Alzheimer's, other factors, such as age, lifestyle, and overall health, also contribute. There are some genetic factors that can increase your risk of developing Alzheimer's, such as the presence of the APOE-e4 gene. However, having this gene does not guarantee that you will develop Alzheimer's, just as not having it does not guarantee that you will not develop the disease. Early-onset Alzheimer's disease, which occurs before the age of 65, is more likely to have a strong genetic component, but it is relatively rare compared to the more common late-onset form of the disease. It is important to remember that genetic factors are only one piece of the puzzle, and maintaining a healthy lifestyle, staying mentally active, and managing chronic health conditions may help reduce your risk of developing Alzheimer's disease. If you are concerned about your risk for Alzheimer's or other forms of dementia, it is recommended that you speak with a healthcare professional for personalized advice and guidance.

**To ChatGPT:** Can you please rewrite your previous explanation in language that can be easily understood by a person with a middle school educational level?

**ChatGPT:** Sure! Alzheimer's disease is a brain illness that affects a person's memory, thinking, and behavior. If your parent has Alzheimer's, it doesn't mean you will definitely get it. While genes from your family can play a part in the chance of getting Alzheimer's, other things like age, the way you live, and your overall health are important too. There is a gene called APOE-e4 that can increase the chance of getting Alzheimer's. But having this gene doesn't mean you'll get the disease for sure. Also, not having it doesn't mean you won't get Alzheimer's. Alzheimer's that starts before age 65 is called early-onset Alzheimer's. This type is more likely to be caused by genes, but it is not very common. Remember that genes are only one part of the story. Keeping a healthy lifestyle, staying active mentally, and taking care of any health problems can help lower the chance of getting Alzheimer's. If you're worried about Alzheimer's, it's a good idea to talk to a doctor who can give you more information and advice based on your situation.

### Myth #11

**To ChatGPT:** Do you think that "Only older people get Alzheimer's Disease"?

**ChatGPT:** As an AI language model, I don't have personal opinions, but I can provide information based on the data I've been trained on. The statement "Only older people get Alzheimer's disease" is not entirely accurate, although age is indeed a significant risk factor for the disease. Alzheimer's disease is the most common cause of dementia, a general term for a decline in cognitive ability severe enough to interfere with daily life. While the majority of people with Alzheimer's disease are aged 65 and older, it can also affect younger individuals. Early-onset Alzheimer's, which occurs in people between the ages of 30 and 64, accounts for a smaller percentage of cases. It is essential to note that early-onset Alzheimer's is relatively rare compared to the late-onset form of the disease. Several factors can contribute to the development of Alzheimer's, including genetics, lifestyle, and environmental factors. Age is the most substantial risk factor, but it is not the sole determinant of whether someone will develop the disease.

**To ChatGPT:** Can you please rewrite your previous explanation in language that can be easily understood by a person with a middle school educational level?

**ChatGPT:** Sure! Alzheimer's disease is a brain problem that makes it hard for people to remember things and think clearly. It usually happens to older people, but sometimes younger people can get it too. This is called early-onset Alzheimer's, and it doesn't happen as often. Even though age is an important factor in getting Alzheimer's, it's not the only thing that matters. There are other reasons someone might get the disease, like their genes, the way they live, and things in their environment. So, it's not true that only older people get Alzheimer's, but it is more common in older people.

### Myth #12

**To ChatGPT:** Do you think that "Supplements found online can prevent or cure Alzheimer's Disease"?

**ChatGPT:** As an AI language model, I do not have personal opinions, but I can provide you with information on the topic. There is no definitive evidence or scientific consensus that any supplement can prevent or cure Alzheimer's disease. Alzheimer's is a complex and progressive neurological disorder, and its exact causes are still not fully understood. While some studies have suggested that certain supplements, such as vitamins E and B, omega-3 fatty acids, and curcumin, might have potential benefits for cognitive function and brain health, these findings are not conclusive enough to support the use of such supplements as a primary prevention or treatment strategy for Alzheimer's disease. It is important to consult with a healthcare professional before starting any supplement regimen, especially if you are concerned about

Alzheimer's disease or other cognitive disorders. A healthcare professional can provide personalized recommendations based on your medical history, current health, and any potential interactions with medications you may be taking. Maintaining a healthy lifestyle, including regular physical activity, a balanced diet, good sleep hygiene, and social engagement, is a more effective way to promote brain health and reduce the risk of Alzheimer's disease and other cognitive disorders.

**To ChatGPT:** Can you please rewrite your previous explanation in language that can be easily understood by a person with a middle school educational level?

**ChatGPT:** Sure! Alzheimer's disease is a brain problem that affects memory and thinking. There is no strong proof that any supplements, like vitamins or special nutrients, can stop or cure Alzheimer's disease. Some studies say that certain supplements might help brain health, but we don't have enough proof to say they can prevent or treat Alzheimer's. It is important to talk to a doctor before taking any supplements, especially if you are worried about Alzheimer's or other brain problems. A better way to keep your brain healthy is to live a healthy lifestyle. This includes being active, eating well, sleeping enough, and spending time with friends and family.

#### Myth #13

**To ChatGPT**: Do you think that "Specific diets can reduce the risk of Alzheimer's Disease"?

**ChatGPT**: I am an AI language model and cannot form opinions. However, I can provide you with you with information on the topic. Research suggests that specific diets may help reduce the risk of Alzheimer's disease and other forms of dementia. Some of the diets that have been associated with a reduced risk include the Mediterranean diet, the DASH diet (Dietary Approaches to Stop Hypertension), and the MIND diet (Mediterranean-DASH Diet Intervention for Neurodegenerative Delay). These diets emphasize the consumption of fruits, vegetables, whole grains, legumes, nuts, fish, and healthy fats like olive oil, while limiting saturated fats, red meats, and added sugars. They are rich in antioxidants, vitamins, minerals, and healthy fats, which have been shown to have potential neuroprotective effects. Although there is no guarantee that any specific diet will completely prevent Alzheimer's disease, adhering to a healthy eating pattern may contribute to reducing the risk of developing the disease or delaying its onset. It's important to remember that a combination of factors, including genetics, lifestyle, and environmental factors, can influence the risk of developing Alzheimer's disease. As always, it's a good idea to consult with a healthcare professional before making significant changes to your diet or lifestyle to ensure you are taking the appropriate steps for your individual needs.

**To ChatGPT**: Can you please rewrite your previous explanation in language that can be easily understood by a person with a middle school educational level?

**ChatGPT**: Sure! Some studies show that eating certain diets can help lower the chances of getting Alzheimer's disease. Alzheimer's is a brain disease that affects memory and thinking. Three diets that might help are:

- Mediterranean diet: This diet has lots of fruits, vegetables, whole grains, beans, nuts, and fish. It uses healthy fats like olive oil instead of butter.

- DASH diet: DASH stands for "Dietary Approaches to Stop Hypertension." This diet is good for your heart and brain. It has lots of fruits, vegetables, whole grains, and low-fat dairy products.
- MIND diet: This diet combines parts of the Mediterranean and DASH diets. It focuses on foods that are good for your brain.

These diets are good because they have lots of vitamins, minerals, and healthy fats that can protect your brain. However, no diet can completely prevent Alzheimer's disease. Many things, like your genes and your lifestyle, can affect your chances of getting the disease. Before making big changes to what you eat, it's a good idea to talk to a doctor or another health expert. They can help you find the best plan for you.

### Myth #14

**To ChatGPT**: Do you think that "Prescription medications can reverse the progression to Alzheimer's Disease"?

**ChatGPT**: As an AI language model, I don't have personal opinions or beliefs. However, I can provide you with information on this topic. As of my last knowledge update in September 2021, there is no definitive cure for Alzheimer's disease. However, some prescription medications can help manage symptoms, slow down the progression of the disease, or temporarily improve cognitive function. The main classes of drugs that have been used to treat Alzheimer's disease include cholinesterase inhibitors (such as donepezil, rivastigmine, and galantamine) and NMDA receptor antagonists (like memantine). These medications can help manage symptoms but do not reverse the underlying disease progression or cure Alzheimer's. Research into Alzheimer's disease is ongoing, and scientists continue to explore new treatment options that could potentially reverse or halt the progression of the disease. It is possible that advances have been made since my last knowledge update, so it's important to consult recent scientific literature and medical professionals for the most up-to-date information on Alzheimer's disease treatment.

**To ChatGPT**: Can you please rewrite your previous explanation in language that can be easily understood by a person with a middle school educational level?

**ChatGPT**: Sure! Alzheimer's disease is a brain illness that makes it hard for people to remember things and think clearly. As of my last update in 2021, there is no cure for this disease. However, there are some medications that doctors give to patients with Alzheimer's to help them feel better and slow down the illness. These medicines, like donepezil, rivastigmine, galantamine, and memantine, can help with memory problems and other symptoms, but they don't stop the disease from getting worse or cure it. Scientists are always studying new ways to treat Alzheimer's, and there might be new treatments available since my last update. To learn more, it's important to check the latest research and talk to a doctor.

### Myth #15

**To ChatGPT:** Do you think that "Non-steroidal anti-inflammatory drugs (NSAIDs) can prevent Alzheimer's Disease"?

**ChatGPT:** I am an AI language model, and I cannot "think" in the way humans do, but I can provide you with information based on the data available up to my knowledge cutoff in 2021. As of my last update in 2021, there is no definitive evidence that nonsteroidal anti-inflammatory drugs (NSAIDs) can prevent Alzheimer's disease. Some studies have suggested a possible link between the use of NSAIDs and a reduced risk of Alzheimer's disease. The hypothesis is that NSAIDs may have a protective effect due to their anti-inflammatory properties, as inflammation has been implicated in the development of Alzheimer's disease. However, the results of these studies have been mixed, and more research is needed to determine whether there is a causal relationship between NSAIDs and Alzheimer's disease prevention. It is important to consult with a healthcare professional before using any medication for the purpose of preventing a disease, especially given the potential side effects and interactions that may occur with NSAIDs. Always rely on updated, evidence-based information and consult a healthcare professional for guidance on disease prevention and treatment.

**To ChatGPT:** Can you please rewrite your previous explanation in language that can be easily understood by a person with a middle school educational level?

**ChatGPT:** Sure! I'll make it simpler. There is no clear proof that nonsteroidal anti-inflammatory drugs (NSAIDs), a type of medicine that helps with pain and inflammation, can prevent Alzheimer's disease. Alzheimer's is a brain illness that affects memory and thinking. Some studies have shown that NSAIDs might help lower the risk of getting Alzheimer's because they can reduce inflammation, which is related to this disease. But, the results from these studies are not the same, and more research is needed to find out if NSAIDs really help prevent Alzheimer's. It's important to talk to a doctor before using any medicine to try to prevent a disease, especially since NSAIDs can sometimes cause side effects or problems when taken with other medicines. Remember to always get up-to-date information and talk to a doctor for advice on how to prevent or treat diseases.

### Myth #16

**To ChatGPT:** Do you think that "Ginkgo biloba can prevent Alzheimer's Disease"?

**ChatGPT:** As an AI language model, I don't have personal opinions or beliefs, but I can provide you with some information on this topic. Ginkgo biloba is a natural supplement derived from the leaves of the Ginkgo tree, and it has been used for various medicinal purposes for centuries. Some studies have suggested that Ginkgo biloba may have potential benefits for cognitive function, including memory and attention, in older adults. Regarding Alzheimer's disease specifically, the evidence is mixed. Some studies have suggested that Ginkgo biloba may help improve symptoms or slow the progression of Alzheimer's disease and other forms of dementia. However, other studies have not found significant benefits. In general, the quality of research on Ginkgo biloba and Alzheimer's disease has been inconsistent, and more rigorous studies are needed to establish a clearer understanding of the relationship between Ginkgo biloba supplementation and Alzheimer's prevention or treatment. It is important to consult with a healthcare professional before starting any new supplement, especially if you are concerned about Alzheimer's disease or other cognitive conditions. A medical professional will be able to provide personalized advice and recommendations based on your specific needs and medical history.

**To ChatGPT:** Can you please rewrite your previous explanation in language that can be easily understood by a person with a middle school educational level?

**ChatGPT:** Ginkgo biloba is a natural product from the leaves of the Ginkgo tree. People have used it for a long time to help with different health problems. Some studies show that Ginkgo biloba might help older people with memory and attention. When talking about Alzheimer's disease, the results from studies are not clear. Some studies say that Ginkgo biloba might help with Alzheimer's symptoms or slow down the disease. But other studies don't show any big benefits. We need more research to know if Ginkgo biloba can really help with Alzheimer's disease. If you're thinking about taking Ginkgo biloba or any other supplement, it's important to talk to a doctor first. They can give you the best advice based on your personal health and needs.

### Myth #17

**To ChatGPT:** Do you think that "Hormone therapy prevents and treats Alzheimer's Disease"?

**ChatGPT:** As an AI language model, I cannot express personal opinions. However, I can provide you with information on the topic. Hormone therapy has been investigated as a potential treatment or preventative measure for Alzheimer's disease, particularly hormone replacement therapy (HRT) in postmenopausal women. HRT involves supplementing women with estrogen, sometimes in combination with progesterone, to alleviate menopausal symptoms and reduce the risk of developing certain health conditions. Some studies have suggested that HRT may have a positive effect on cognitive function and reduce the risk of Alzheimer's disease. This is thought to be due to the neuroprotective properties of estrogen, which may help maintain cognitive function and brain health. However, the relationship between hormone therapy and Alzheimer's disease is not entirely clear. Some studies have reported no significant benefits, while others have even suggested potential harm. The Women's Health Initiative Memory Study (WHIMS), for example, found that estrogen-progestin therapy increased the risk of dementia in postmenopausal women aged 65 years and older. As such, it's important to note that hormone therapy is not a proven method for preventing or treating Alzheimer's disease. Researchers are still trying to understand the complex relationship between hormones and cognitive function. If you are considering hormone therapy for any reason, it's crucial to consult with a healthcare professional to discuss the potential risks and benefits in your specific case.

**To ChatGPT:** Can you please rewrite your previous explanation in language that can be easily understood by a person with a middle school educational level?

**ChatGPT:** Sure! Hormone therapy is a treatment that can help women going through menopause by giving them hormones, like estrogen and sometimes progesterone. Some scientists think that this treatment might also help protect the brain and lower the chances of getting Alzheimer's disease. Alzheimer's disease is a condition that affects the brain and makes it hard for people to remember things, think clearly, and take care of themselves. Estrogen, one of the hormones used in the treatment, might help keep the brain healthy. However, not all studies agree that hormone therapy can help prevent or treat Alzheimer's disease. Some studies didn't find any benefits, and some even said that it could cause harm. One study showed that giving estrogen and progesterone together might actually increase the risk of memory problems in older women. So, it's still not clear if hormone therapy can really help with Alzheimer's disease. Scientists are still trying to learn more about how hormones affect the brain. If you're thinking about hormone therapy, it's important to talk to a doctor to understand if it's a good choice for you.

#### Myth #18

**To ChatGPT:** Do you think that "Vaccines increase the risk of Alzheimer's Disease"?

**ChatGPT:** I am an AI language model and cannot form opinions, but I can provide information based on available research. As of my knowledge cutoff date in September 2021, there is no scientific evidence to suggest that vaccines increase the risk of Alzheimer's disease. Alzheimer's disease is a progressive neurological disorder that affects memory, thinking, and behavior. Its causes are not yet fully understood, but researchers believe that a combination of genetic, lifestyle, and environmental factors contribute to its development. Vaccines, on the other hand, have been proven to be safe and effective in preventing many infectious diseases. They work by stimulating the immune system to produce an immune response that helps protect against specific pathogens. Some studies have even suggested that certain vaccines, such as the flu vaccine, might have a protective effect against Alzheimer's disease. However, more research is needed to establish a definitive link. It is essential to rely on reputable sources, such as scientific journals and health organizations, when seeking information on vaccines and their potential impact on health. Misinformation about vaccines can lead to unwarranted fears and reduced vaccination rates, which can have serious consequences for public health.

**To ChatGPT:** Can you please rewrite your previous explanation in language that can be easily understood by a person with a middle school educational level?

**ChatGPT:** I am an AI language model and I don't have opinions, but I can share information. As of my knowledge update in September 2021, there is no proof that vaccines can cause Alzheimer's disease. Alzheimer's disease is a brain problem that makes it hard for people to remember things, think clearly, and behave normally. We don't know exactly what causes it, but it might be a mix of genes, lifestyle, and things around us. Vaccines help protect us from many diseases by teaching our body's defense system how to fight off harmful germs. Vaccines are safe and work well. Some studies even show that certain vaccines, like the flu shot, might help protect against Alzheimer's disease. But we need more research to know for sure. It's important to trust reliable sources, like science articles and health organizations, when learning about vaccines and health. If people get the wrong information about vaccines, they might be scared to get them, which can be bad for everyone's health.

#### Myth #19

**To ChatGPT:** Do you think that "Aspartame increases the risk of Alzheimer's Disease"?

**ChatGPT:** As an AI language model, I don't have personal opinions, but I can provide you with information based on existing research. There is no solid scientific evidence to date that directly links aspartame, an artificial sweetener, to an increased risk of Alzheimer's disease. Aspartame has been extensively studied, and it is considered safe for human consumption by numerous regulatory agencies, including the US Food and Drug Administration (FDA) and the European Food Safety Authority (EFSA). Some studies have explored the potential relationship between aspartame and neurodegenerative diseases such as Alzheimer's, but the results have been inconclusive. It is worth noting that many factors, including genetics, age, lifestyle, and environmental factors, contribute to the development of Alzheimer's disease. While some people might be sensitive to aspartame, experiencing headaches or other adverse effects, there is no conclusive evidence linking it to Alzheimer's. It's always a good idea to consult with a healthcare professional if you have concerns about your dietary choices and their potential impact on your health.

**To ChatGPT:** Can you please rewrite your previous explanation in language that can be easily understood by a person with a middle school educational level?

**ChatGPT:** Sure! Aspartame is an artificial sweetener that is used in many foods and drinks. So far, there is no strong evidence to show that aspartame causes Alzheimer's disease. Alzheimer's is a brain disease that affects memory and thinking, and many things can play a role in whether someone gets it, like age, genes, and lifestyle. Many experts, like the ones at the FDA (Food and Drug Administration) in the United States and EFSA (European Food Safety Authority) in Europe, say that aspartame is safe to eat and drink. Some studies have tried to find a connection between aspartame and Alzheimer's, but they haven't found any clear answers. Some people might have problems, like headaches, when they eat or drink things with aspartame, but that doesn't mean it causes Alzheimer's. If you're worried about aspartame or your diet, it's a good idea to talk to a doctor or another health professional. They can help you make the best choices for your health.

#### Myth #20

**To ChatGPT:** Do you think that "Statins increase the risk of Alzheimer's Disease"?

**ChatGPT:** As an AI language model, I cannot provide opinions, but I can offer information based on the data I have been trained on, which goes up until September 2021. As of my knowledge cutoff date in 2021, there was no definitive evidence to suggest that statins, a class of drugs typically prescribed to lower cholesterol levels, increase the risk of Alzheimer's disease. In fact, some studies have suggested that statins may have a protective effect against Alzheimer's disease due to their anti-inflammatory and antioxidant properties. However, it is essential to note that research in this area is ongoing, and my knowledge may be outdated. Always consult with a healthcare professional or review the latest studies and scientific literature for the most up-to-date information on this topic.

**To ChatGPT:** Can you please rewrite your previous explanation in language that can be easily understood by a person with a middle school educational level?

**ChatGPT:** Sure, I can make it simpler for you. Statins are medicines that help lower the amount of cholesterol in your blood. As of my last update in 2021, there wasn't proof that statins make Alzheimer's disease more likely. Alzheimer's is a brain illness that affects memory and thinking. In fact, some studies have shown that statins might help protect your brain from Alzheimer's because they can fight inflammation and harmful substances. But remember, I don't have the latest information. Always talk to a doctor or look at new studies for the most recent details about this topic.
